# Risk Assessment and Mitigation of Airborne Disease Transmission in Orchestral Wind Instrument Performance

**DOI:** 10.1101/2020.12.23.20248652

**Authors:** Aliza Abraham, Ruichen He, Siyao Shao, S. Santosh Kumar, Changchang Wang, Buyu Guo, Maximilian Trifonov, Rafael Grazzini Placucci, Mele Willis, Jiarong Hong

**Affiliations:** Department of Mechanical Engineering, University of Minnesota, Minneapolis, MN, USA 55414; Saint Anthony Falls Laboratory, University of Minnesota, Minneapolis, MN, USA 55414; Minnesota Orchestra, 1111 Nicollet Mall, Minneapolis, MN 55403

## Abstract

There has been growing concern about high risk of airborne infection during wind instrument performance as the COVID-19 pandemic evolves. In collaboration with 16 musicians from the Minnesota Orchestra, we employ multiple experimental and numerical techniques to quantify the airflow and aerosol concentration emitted from ten wind instruments under realistic performance conditions. For all instruments, the extent of the flow and aerosol influence zones are limited to 30 cm. Further away, the thermal plume generated by the human body is the dominant source of flow. Flow and aerosol concentration vary in response to changes in music amplitude, pitch, and note duration, depending on playing technique and instrument geometry. Covering the trumpet bell with speaker cloth and placing filters above the instrument outlet can substantially reduce the aerosol concentration. Our findings indicate that with appropriate risk mitigation strategies, musical instrument performance can be conducted with low risk of airborne disease transmission.

---

Growing evidence indicates the potential for the transmission of COVID-19 through aerosols, i.e. small airborne particles (typically < 5 μm in diameter), produced by even normal human respiratory behaviors such as breathing and speaking^1–4^. Viruses attach to these aerosols, which can stay suspended in the air for hours and accumulate, particularly in indoor spaces, posing significant infection risk to individuals who inhale them^5–9^. Masks covering the nose and mouth have been widely adopted as a way to minimize inhalation and emission of aerosols^10–12^. However, such preventive measures are difficult to implement during some musical activities such as singing and wind instrument play. Without additional precautions to mitigate risks during these activities, increased spread of infection and loss of life can result. Recently, choirs from Washington state^13^ and Amsterdam^14^ experienced tragedies when its members were infected with COVID-19 during regular rehearsals and performances, resulting in multiple deaths, even while following standard social distancing guidelines. These devastating experiences exacerbate concerns about practice and performance of brass and woodwind instruments, where aerosols may be produced at higher concentrations and spread farther distances. However, to date, very few studies have investigated aerosol production and transport by musical instruments. One recent study provided a detailed investigation of aerosol generation from multiple different wind instruments, revealing large variability across instruments, including some that produce significantly more aerosols than breathing and speaking^15^. However, to accurately assess the risk of COVID-19 transmission during orchestra rehearsal and performance, the aerosol transport must be investigated along with their production. Only a handful of preliminary studies have been conducted to quantify the air flow generated by different instruments. Becher et al.^16^ used schlieren imaging to visualize the flow emitted from multiple instruments, but did not quantify the velocity or measure the aerosol content of the flow. Kähler and Hain^17^ showed that woodwind instruments, particularly the flute, influence the flow in a much larger region than brass instruments. Spahn and Richter^18^ provided guidelines for musician spacing based on air flow measurements conducted at the Bamberg Symphony Orchestra, suggesting that 2 m spacing is still sufficient as no instrument-induced air movement is detected at this distance. No flow measurements at closer distances were provided. However, the aforementioned studies are not peer reviewed, and none quantify the spread of aerosols from the instruments, nor the effect of convective flow from the body heat of the musicians. Furthermore, the flow visualizations presented by Kähler and Hain^17^ and Spahn and Richter^18^ were conducted in laboratory settings where the ventilation is drastically different from a large performance hall. To provide a more realistic assessment of the risk associated with musical performance, we collaborate with 16 musicians from the Minnesota Orchestra to systematically examine aerosol generation and transport from 10 woodwind and brass instruments (Supplementary Fig. 1) at the Minnesota Orchestra Hall. We find that the influence zone is smaller than previously reported, as natural convection from the bodies of the musicians enhances mixing between the flow expelled from the instrument and the ambient air. The aerosol concentration also disperses rapidly, dropping to background levels by 30 cm. Based on these findings, we propose and evaluate multiple mitigation techniques to avoid aerosol accumulation within the orchestra hall such as masks for the instrument outlets and air filters placed above the musicians to capitalize on the upward motion of the human thermal plume. Our findings can be further generalized to other woodwind and brass instruments not included in the present study and for the safe arrangement of different musical performance settings.

## Results and Discussion

### Flow influence zone

The flow influence zone of each instrument is first evaluated using a schlieren flow measurement method. This technique enables the visualization of air density changes caused by temperature gradients, revealing air flow emitted from a person’s breath (warmer than ambient air). When a musician blows into their instrument, the plume of air emitted from the instrument outlet is clearly visible, and the velocity of the flow can be quantified (Fig. 1a). Using the outlet velocity and the length of the disturbed region of air (i.e., the region where the velocity is greater than the background velocity caused by the ventilation and the thermal plume near the body), the strength and extent of the influence zone of each instrument is quantified (Fig. 1b). The two instruments with the highest outlet velocities are flute and clarinet. Both of these woodwind instruments have relatively narrow outlets (Supplementary Table 2), which cause a given volume of air to move faster at the exit than in instruments with wider bells. The bass clarinet, for example, has a similar mouthpiece configuration to the clarinet, but its wider bell decelerates the flow at the outlet. The bassoon and oboe also have narrow bells, but their double reed mouthpieces allow a lower volume of air into the instrument compared to single-reed and air jet instruments^19^. Still, between the bassoon and the oboe, the reed opening on an oboe is narrower than that on a bassoon, causing increased flow resistance and lower outlet velocity. Furthermore, the long upward-oriented vertical section of the bassoon induces flow acceleration due to buoyancy, further increasing the velocity at the outlet. Among the brass instruments, the trumpet has the smallest bell size and correspondingly the highest outlet velocity. The French horn has the lowest outlet velocity because the instrument is played with the musician’s hand in the bell, which interferes with the flow at the outlet.

**Figure 1.**
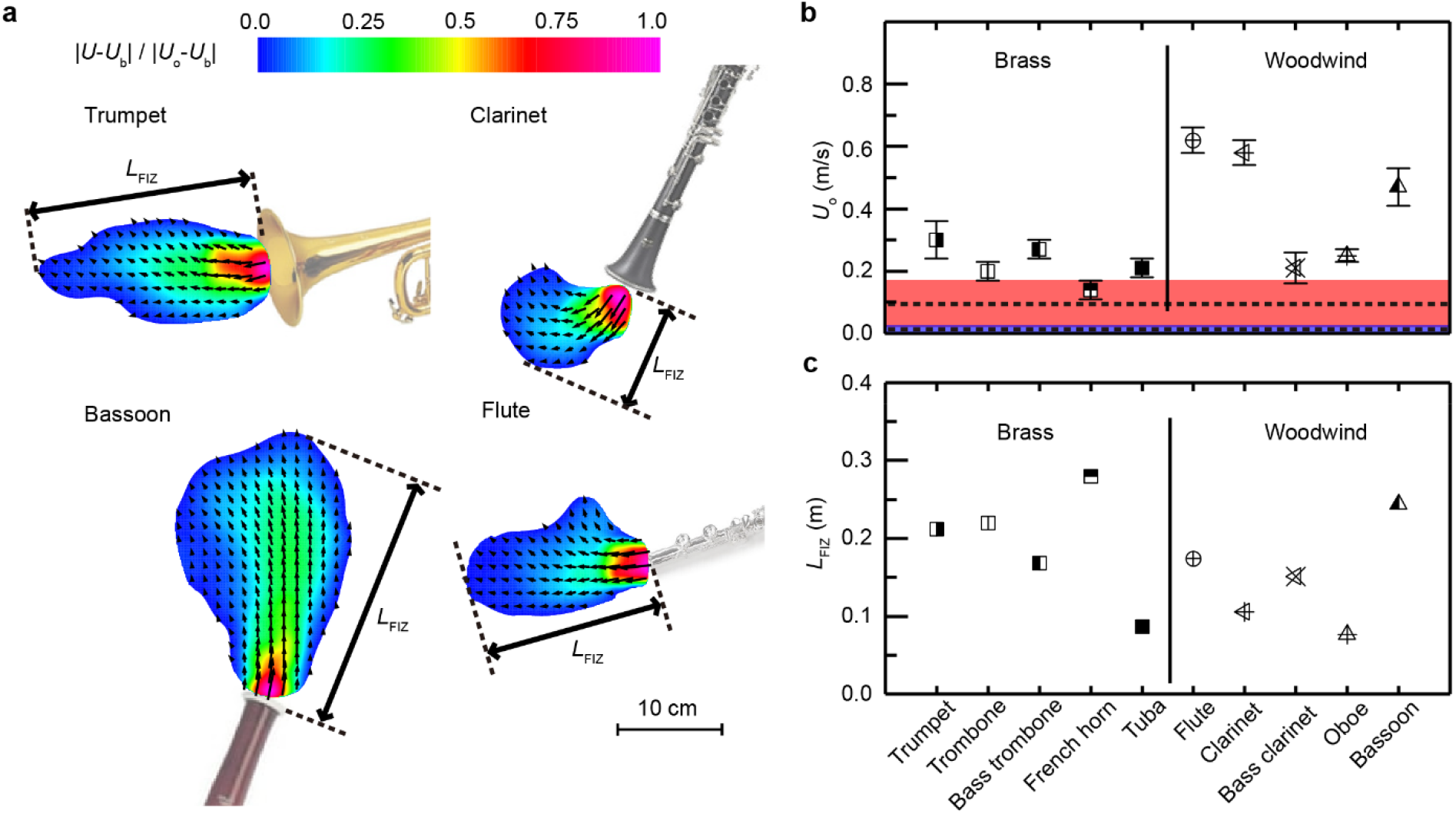
Flow influence zone. The region influenced by the flow of each instrument with (**a**) examples shown for trumpet, clarinet, bassoon, and flute, and influence zone parameters, including (**b**) outlet velocity (*U*_o_), thermal plume (mean indicated by dashed horizontal line and standard deviation represented by red shaded area, see Supplementary Fig. 8) and ventilation velocity (mean indicated by dashed horizontal line and standard deviation represented by blue shaded area, see Supplementary Fig. 6), and (**c**) flow influence zone length (*L*_FIZ_) for all instruments. *L*_FIZ_ is determined by the maximum extent of the influence zone in the direction of the outlet flow. Note that the air flow from the instruments is intermittent depending on the breathing pattern and music performed. The error bars for *U*_o_ indicate the standard deviation of the outlet velocity during the period of consistent flow used to generate the velocity field for each instrument. The velocity fields of the instruments not shown here can be found in Supplementary Fig. 2. For the trombone, bass trombone, and French horn, *U*_o_ is determined using anemometer data, as the musician’s hand in or near the bell interferes with near-field flow measurements using schlieren data. Sample enhanced schlieren videos can be found in Supplementary Videos 1-5.

The lengths of the influence zones of all instruments are generally proportional to the maximum outlet velocities, with some exceptions. First, the instruments with larger bell sizes, i.e., the brass instruments, have longer influence zones than that expected based on velocity alone, as a wider jet of air will extend farther than a narrow jet with the same velocity^20^. Second, the direction of the outlet relative to the thermal body plume affects the influence zone length. Instruments with outlets that point upwards have longer influence zones (e.g., bassoon, bass clarinet), as the thermal plume works constructively with the instrument flow to lengthen the influence zone (Supplementary Fig. 2). On the other hand, downward-directed instruments (e.g., clarinet, oboe) have shorter influence zones due to the destructive interaction between the instrument flow and the thermal plume. In particular, oboists must employ a special breathing technique to account for the low volume of air allowed into the instrument^19^, resulting in the very confined influence zone of the oboe (Supplementary Fig. 2). Because the French horn is played with the musician’s hand inside the bell, the thermal plume from the arm extends the influence zone beyond that of any other instrument.

In general, all instruments exhibit a smaller influence zone than that reported in preliminary studies^16,17^ which suggested the flow from certain instruments such as the flute and the bassoon could influence the air greater than 1 m away from the musician. The influence zone observed in the current study is confined to 30 cm for all instruments. This discrepancy is attributed to the difference in room sizes between the previous studies conducted in laboratory settings and the current study conducted in the much larger Orchestra Hall. In smaller rooms, ventilation tends to be stronger than in larger rooms such as concert halls^18,21^, which could cause the influence zone to extend further in the horizontal direction. In Orchestra Hall, the thermal plume caused by the temperature difference between the body of the musician and the ambient air dominates over the weaker ventilation, directing the influence zone upwards and enhancing mixing. At its maximum, the velocity of the thermal plume is comparable to that of the outlet flow for some instruments (Fig. 1b). Considering the aforementioned effects, the influence zone of all instruments is comparable to that of normal breathing^22^, suggesting no additional spacing between musicians beyond the typically recommended 6 feet is required during musical instrument play.

### Aerosol influence zone

We next quantify the distribution of aerosols emitted by each instrument, and the extent of their influence. Using an aerodynamic particle sizer (APS), we measure the concentration of aerosols at the instrument outlet and at multiple locations in each direction parallel and perpendicular to the outlet flow. We supplement these point measurements with CFD simulations initialized with experimental concentration and flow data. The region around the outlet of each instrument in which the aerosol concentration is greater than the background concentration in Orchestra Hall (1.2 particles/s or 14.4 particles/L) is defined as the aerosol influence zone (Fig. 2a). The extent of the aerosol influence zone for each instrument (Fig. 2b) depends on both the flow influence zone and the aerosol concentration at the outlet. For example, the trumpet exhibits the longest aerosol influence zone due to its relatively long flow influence zone and its high outlet concentration (consistent with He et al.^15^). On the other hand, the bassoon generates a flow influence zone longer than that of the trumpet, but its aerosol influence zone is significantly smaller due to its lower outlet concentration. Note that French horn (played with the musician’s hand in the bell) and tuba are not included because they both emit aerosol concentrations at the background level based on APS measurements. Additionally, the influence zone of the flute embouchure is not simulated due to the complexity of the flow from the mouth over the mouthpiece. However, the experimental measurements (included in Supplementary Fig. 4) indicate the aerosol concentration in this region is very low, consistent with the findings of He et al.^15^.

**Figure 2.**
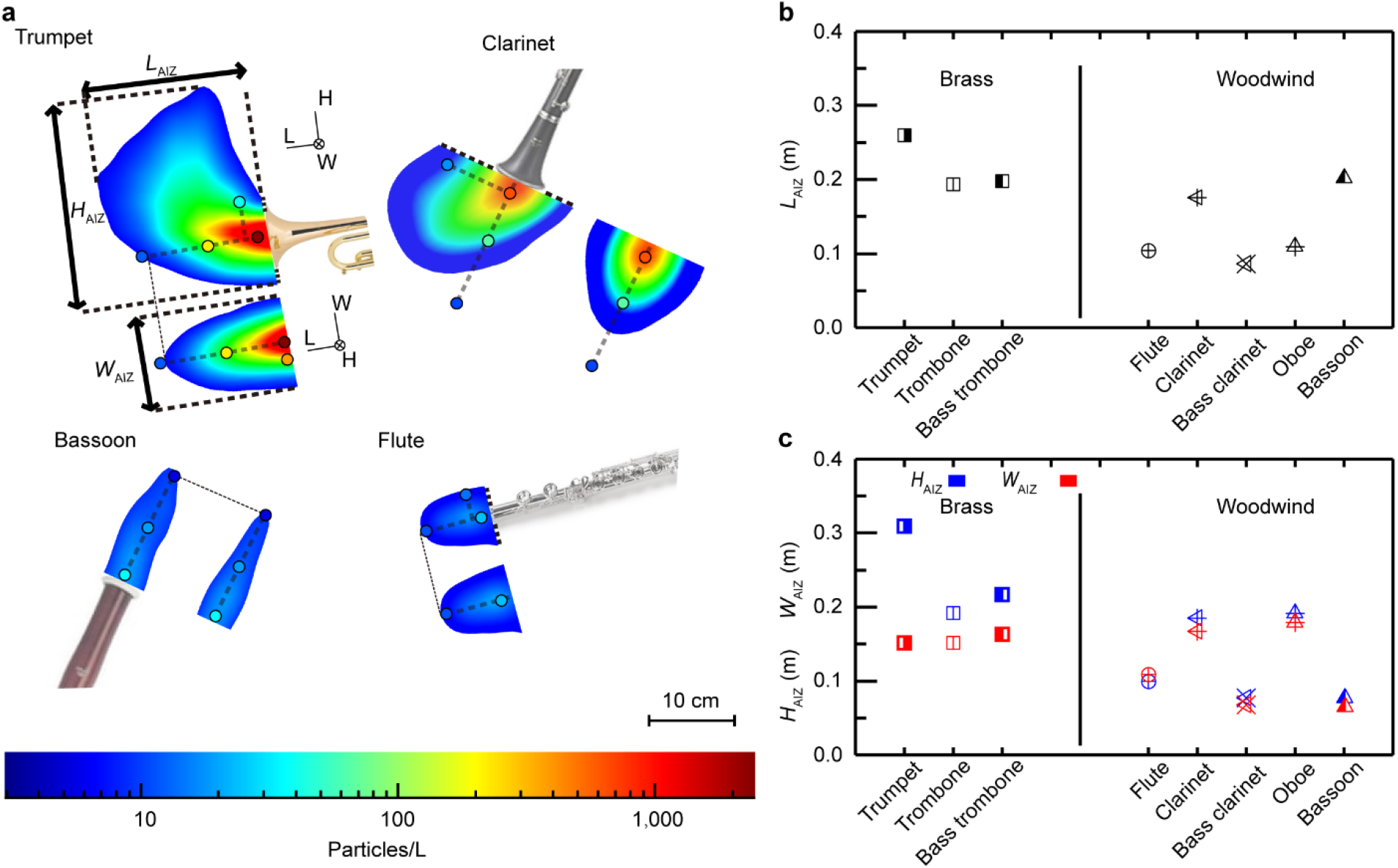
Aerosol influence zone. The particle concentration throughout cross-sections of the instrument aerosol influence zone with (**a**) examples shown for trumpet, clarinet, bassoon, and flute, including dots showing experimental measurement locations and values; (**b**) the length of the aerosol influence zones (*L*_AIZ_); and (**c**) the width (*W*_AIZ_) and height (*H*_AIZ_) of the aerosol influence zones all instruments. The aerosol influence zone is defined as the region around the outlet of each instrument in which the aerosol concentration is greater than the background concentration in Orchestra Hall (1.2 particles/s or 14.4 particles/L), and *L*_AIZ_, *H*_AIZ_, and *W*_AIZ_ are the maximum extents of the influence zone in the directions parallel and perpendicular to the outlet flow, respectively. The aerosol influence zones of instruments not shown here can be found in Supplementary Fig. 3. Note that French horn (played with the musician’s hand in the bell) and tuba are not included because they both emit aerosol concentrations at the background level based on APS measurements. A sample video of the aerosol transport simulation for the trumpet can be found in Supplementary Video 6.

The aerosol influence zone width in the lateral direction and height in the vertical direction are also quantified. For the brass instruments, the widths increase with increasing bell size. The height of the trumpet aerosol influence zone is significantly larger than any of the other instruments, which is due to the fact that it emits the most aerosols. The emitted aerosols are not fully dispersed before the human body-induced thermal plume begins to transport them upwards, extending the height of the aerosol influence zone. Among the woodwinds, the clarinet and oboe have the largest influence zone widths and heights, as they emit the more aerosols than the other three woodwinds (see Supplementary Fig. 3 and He et al.^15^).

Though the aerosol influence zones vary, all instruments exhibit aerosol concentrations that reach the background level within 30 cm of the outlet, and well within the 6-foot spacing frequently recommended for social distancing. However, several instruments produce significantly more aerosols than breathing, exacerbating the risk of particle accumulation around the musicians unless appropriate mitigation strategies are implemented.

### Flow and aerosol correlation with music amplitude, pitch, and note duration

We observe some interesting trends by looking at the changes in music amplitude, pitch, and note duration, and the corresponding changes in flow speed and aerosol concentration emitted from the instruments (Fig. 3a). For example, the flow speed from the trumpet and trombone exhibit a clear relationship with music amplitude (Fig. 3b), while the woodwind instruments do not (Supplementary Fig. 5), due to the larger flow rate variations required for amplitude changes in brass instruments relative to woodwinds^19^. The flow from the bass trombone is not strongly correlated with amplitude either, which we attribute to the larger air requirement of bass trombones due to their wider mouthpiece, bore, and bell^23^, making it more difficult to increase the amplitude by further increasing the flow rate, or to variability in playing style. The bass trombonist from the current study noted that the sound from the instrument can be loud and penetrating, so he tries to “warm” the sound, which could be indicative of a slower air column, even at higher amplitudes. The brass instruments also exhibit some relationships with pitch, which is exhibited most clearly in the bass trombone where the flow rate is not influenced by amplitude. The flow rate and pitch are inversely related, as lower notes require a more open embouchure which allows more flow into the instrument^24–26^. This trend weakens moving to trombone and trumpet due to increasing correlations between pitch and amplitude in the music (i.e., higher notes are played louder) which mask the relationship between pitch and flow speed with the stronger relationship between amplitude and flow speed. Furthermore, the bass trombone requires the most open embouchure to generate the necessary air flow, further strengthening the correlation^23^. Among the woodwinds, the bassoon shows a strong inverse correlation between flow speed and pitch (Fig. 3a,b), caused by the fact that more tone holes are covered for lower notes, pushing more air out of the bell. While this method of changing the pitch is used for all woodwinds, the bassoon demonstrates the relationship most clearly because of the tone holes located near the bottom of the instrument, just above a 180° turn. The centrifugal effects of the 180° bend generate a secondary flow called Dean flow that pushes the air towards the outer edge of the turn, and persists to several times the pipe diameter downstream of the bend^27–29^ (see Supplementary Fig. 7). In the other woodwind instruments, the tone holes are all located along a straight bore, so no secondary flow develops to direct air out of the tone holes.

**Figure 3.**
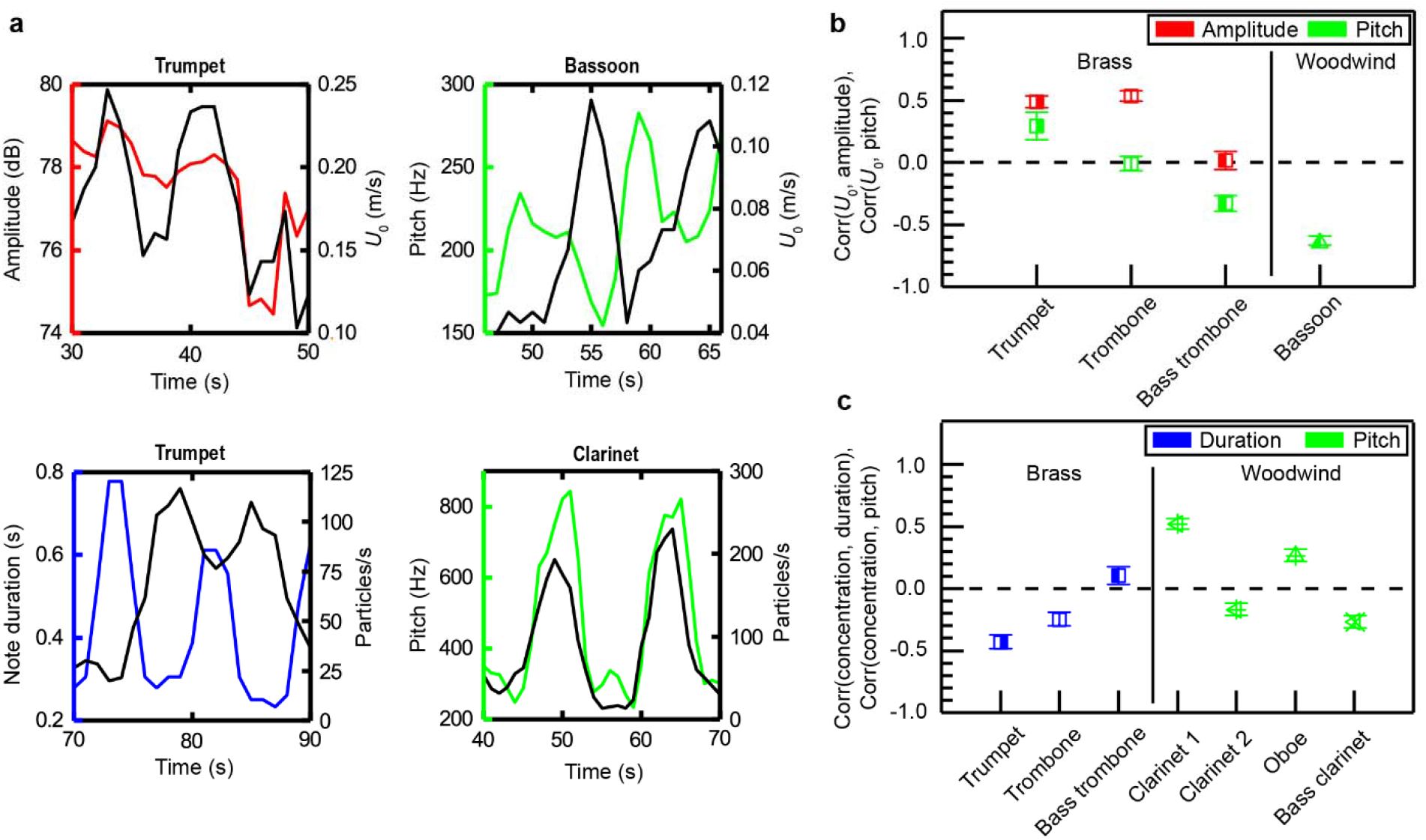
Correlations between pitch, amplitude, note duration, flow speed and aerosol concentration. (**a**) Sample time sequences highlighting the significant correlations between music amplitude and flow speed at the outlet (*U*_*o*_) for the trumpet, pitch and *U*_*o*_ for the bassoon, note duration and aerosol concentration for the trumpet, and pitch and aerosol concentration for the first of the two clarinet players who participated in the experiment. (**b**) Correlation coefficients between *U*_*o*_ and music amplitude, Corr (*U*_*o*_, amplitude), and *U*_*o*_ and pitch, Corr (*U*_*o*_, pitch), for selected instruments. (**c**) Correlation coefficients between aerosol concentration and music duration, Corr concentration, duration), and concentration and pitch, Corr concentration, pitch), for selected instruments. The error bars represent the probable error of each correlation coefficient. All correlation coefficients for all instruments are shown in Supplementary Fig. 5. Sample videos showing time series of all variables can be found in Supplementary Videos 7-10.

The trumpet also shows a strong inverse correlation between aerosol concentration and note duration, indicating more particles are generated when the notes change quickly (Fig. 3c). We attribute this relationship to the fact that brass musicians use the motion of their tongues to separate notes (referred to as articulation). Previous investigations into particle generation during speaking have shown that plosive consonants (e.g., ‘B’, ‘T’, ‘D’, ‘P’) generate more particles than other sounds as the tongue or lips temporarily block the vocal tract, causing the pressure to build up and suddenly release^30,31^. Similarly, brass musicians enunciate a ‘D’ or ‘T’ sound to articulate their music^26^, thereby generating particle-laden bursts with each note. The relationship between note duration and aerosol concentration is clearest for the trumpet and decays in strength for trombone and bass trombone due to the increase in bore length (Supplementary Table 2) and number of bends in the tubing (Supplementary Fig. 1) where particles deposit. This phenomenon does not lead to a significant relationship between velocity and note duration because a series of puffs quickly approaches the typical behavior of a conical jet caused by continuous flow^30^. Among the woodwinds, the relationship between pitch and aerosol concentration for the clarinet is particularly interesting. Two clarinet players participated in the experiment, and the first (highlighted in Fig. 3a) shows a strong correlation, while the second shows almost none (Fig. 3c). We attribute this discrepancy to the musical pieces selected by each musician: The first clarinet performed a piece that included frequent notes in the altissimo range, while the second never entered this range. Previous studies have shown that the vocal tract in clarinet players contributes more for notes in the altissimo range^32^, and the vocal tract is one of the main sources of aerosols^33^. We further support this explanation using the correlations between pitch and particle concentration for other woodwinds. Both oboes show some correlation, and they both performed pieces with occasional altissimo notes, while the bass clarinet shows little correlation and, like the second clarinet, performed a piece that never entered the altissimo range. The bassoon and flute do not generate enough particles to calculate correlation coefficients with confidence. Note that no instruments show a significant correlation between particle concentration and music amplitude (Supplementary Fig. 5), which is consistent with the result of a laboratory study that showed the particle concentrations at different dynamic levels were on the same order of magnitude for all instruments^15^.

### Risk mitigation strategies

We explore multiple strategies for minimizing the risk of exposure to the virus via aerosols generated during instrument play. First, inspired by masks used to cover the nose and mouth, we test the effectiveness of a mask covering the outlet of the instrument. We use Mellotone Acoustic Fabric made by Wendell Fabrics for the mask due to its high sound transmissibility. Using a photograph of the speaker cloth at high magnification, we calculate the porosity of the mask as ϕ = 0.19 (Fig. 4a). We then compare the particle concentration at the outlet of one of the trumpets (the instrument that emits the most particles) without a mask, and with one, two, and three layers of speaker cloth. The reduction in particle concentration with the number of layers increases as a decaying exponential that approaches 1 asymptotically, indicating diminishing returns with each layer (Fig. 4b). With three layers, the particle concentration is reduced to 10% of its initial value, bringing it to a concentration level typical of particle emission from speaking^15^.

**Figure 4.**
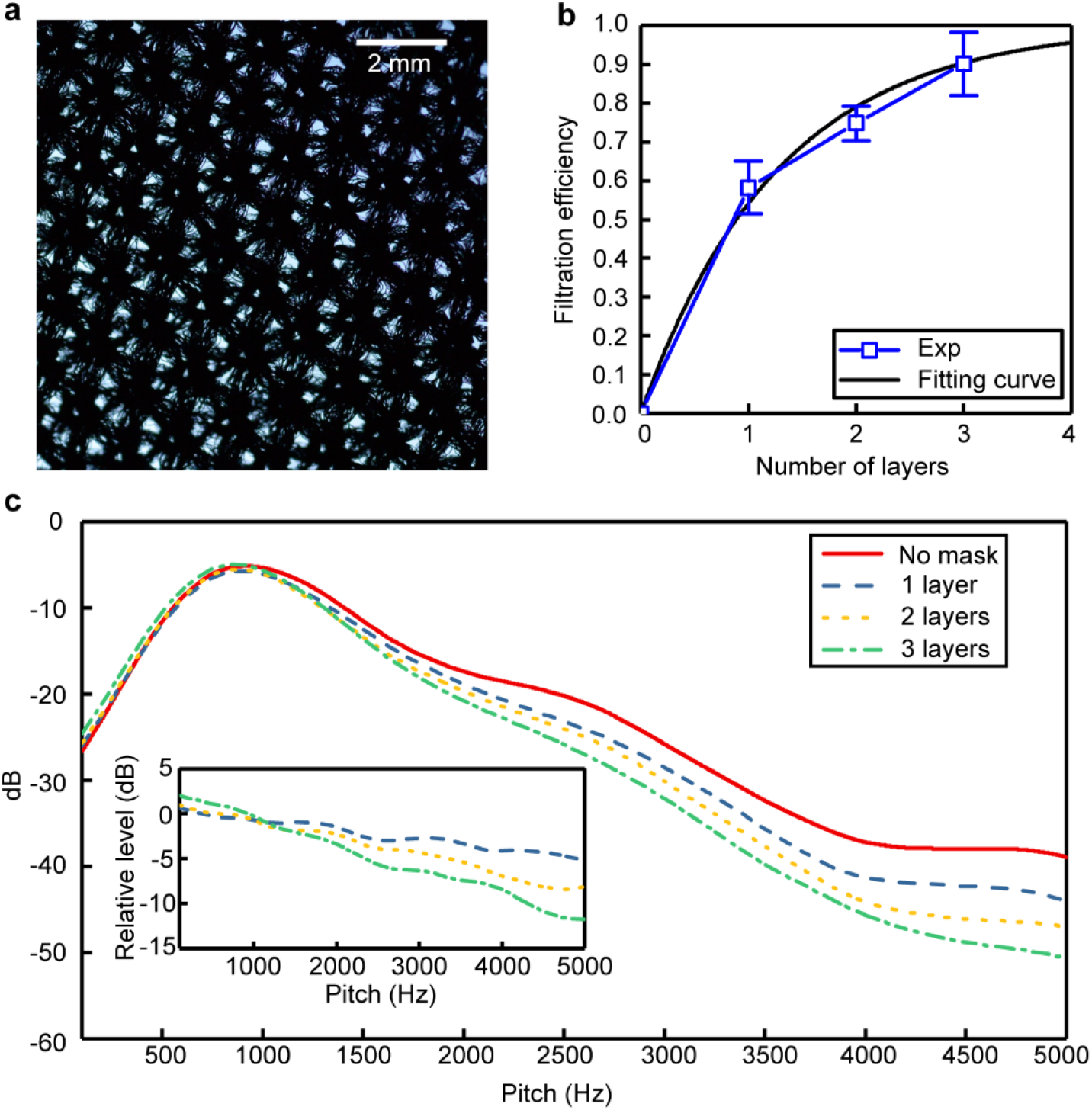
Mask effects. The effect of covering the trumpet bell with one, two, and three layers of speaker cloth as a mask, including (**a**) a magnified sample of the speaker cloth tested with porosity *ϕ = 0*.*19* and its influence on (**b**) outlet particle concentration, including a fit of the form *y* = 1 – exp (*-ax*), where *a* = 0.78, and (**c**) decibel level of the music, considering pitches between 100 Hz and 5000 Hz. The inset shows the decibel level relative to the music performed without a mask.

On the other hand, we record the audio of the musician performing his selected musical piece to assess the reduction in sound quality caused by the additional resistance of the mask material. In general, increasing the number of mask layers decreases decibel level of the music, especially at frequencies above 1000 Hz (Fig. 4c), similar to the observed effects of face masks on speech signals^34^. The trumpet primarily plays notes with fundamental frequencies between 100 Hz and 1000 Hz, but the higher-frequency harmonics and partials contribute significantly up to several thousand Hz, depending on the amplitude^35^. These higher frequencies have an important impact on the timbre (tone quality) of the music. At frequencies below 1000 Hz, the decibel level appears to increase with the number of mask levels, presumably because the musician was compensating for the additional resistance by playing louder. The musician described the feeling of playing with the mask layers as follows: One layer was acceptable, two layers was like playing with a mute (typically used for jazz music but not classical), and three layers was “close to unplayable.” Based on this quantitative and qualitative assessment, we recommend instrument masks should avoid layering if possible. Instead they should be designed using a single layer of low-porosity fabric.

Another popular risk-mitigation strategy is the use of air filters to prevent aerosol accumulation within an indoor space. We simulated filters at different locations relative to the same trumpet player under different conditions during two minutes of music play (Fig. 5). When there is no filter, the majority of aerosols emitted by the musician are transported vertically by the human body-induced thermal plume after a short distance away from the musician, and a region with *I*_risk_ > 10 is formed within 1 m of the musician, where *I*_risk_ indicates the risk of encountering virus-containing particles at a given location^22^ (Fig. 5a). A comparison between a filter placed in front of the musician (Fig. 5b) and a filter placed above (Fig. 5c) shows that the latter placement is drastically more effective at removing aerosols emitted by the instrument. The placement on the floor in front of the musician removes only 3% of the aerosols while the placement above removes 90%, resulting in a reduction in the extent of the region with high *I*_risk_ to 80 cm. The difference between the two configurations is due to the relatively weak flow generated by the filter (set to the lowest flow rate to minimize the amount of performance-disrupting sound generated), which relies instead on the ambient flow to direct particles into the filter. As demonstrated in Fig. 5a, the upward-moving thermal plume generated by the body is the dominant source of flow outside of the immediate instrument influence zone (*L*_*FIZ*_ < 30 cm). Therefore, placing the filter above the musician capitalizes on this upward flow to direct aerosols into the filter most effectively. Furthermore, the filter placed in front of the musician actually increases the spread of aerosols, as evidenced by the increased /_risk_ scatter in the plane at mouth height (bottom panels of Fig. 5b versus Fig. 5a) and the increased width of the high-*I*_risk_ region on the plane behind the musician (middle panels of Fig. 5b versus Fig. 5a). This detrimental effect is due to the local disturbance to the flow caused by the filter.

**Figure 5.**
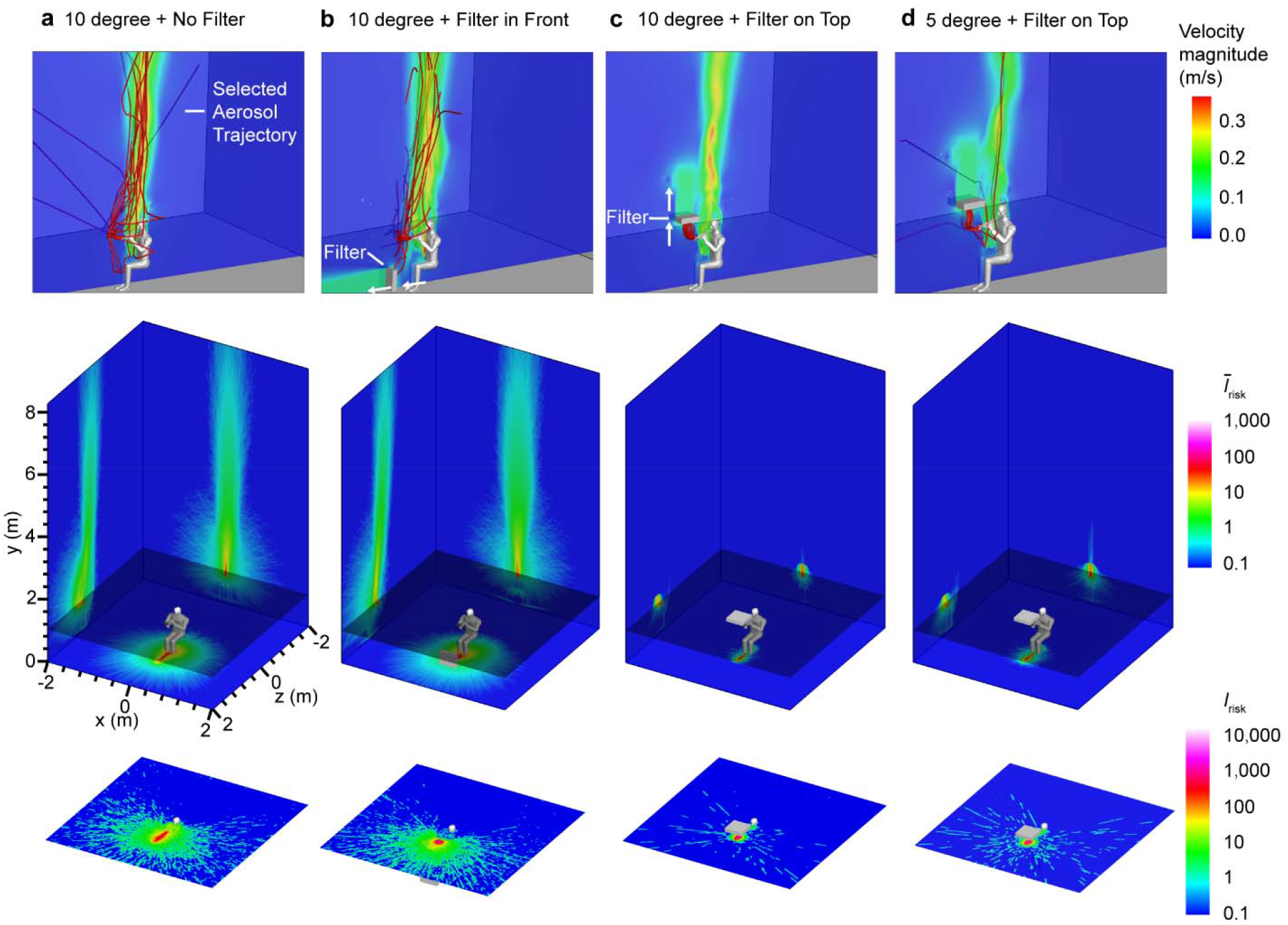
Filtration effects. The air flow velocity (top) and risk of a person encountering virus-containing particles at a given location (*I* _risk_)^22^ within a volume (middle) and in a cross-sectional plane at mouth height (bottom) due to aerosols emitted by a musician with (**a**) no filtration, (**b**) an air filter placed in front of the instrument outlet, and (**c**) an air filter placed above the instrument outlet with an ambient temperature of 20°C (10°C temperature difference with the human body). (**d**) The *I*_risk_ with an air filter placed in front of the instrument outlet and an ambient temperature of 25°C (5°C temperature difference with the human body). The top panels show sample aerosol trajectories with the filter flow direction indicated by white arrows. The middle panels show *Ī*_risk_ on each wall, which is the spatial average of *I*_risk_ along the *x, y*, and *z* directions, whereas the bottom panels show a cross-section *I*_risk_ at the human mouth height. Note that the simulated domain is 8 × 8 × 10 m^3^, but a smaller region is shown here to highlight the area around the musician. Corresponding videos for each case can be found in Supplementary Videos 11-14.

The thermal plume is generated by the temperature difference between the human body and the surroundings. This plume can be strengthened by increasing the temperature difference, i.e., reducing the ambient temperature. We investigate this effect by comparing a 10°C temperature difference to a 5°C temperature difference between the ambient air and the human body temperature. These temperatures were selected to model the typical stage temperature at Orchestra Hall during a performance and a slightly reduced temperature to enhance the thermal plume. The 10°C temperature difference strengthens the thermal plume to generate an improvement in filtration efficiency of 8%, from 83% to 90% of the aerosols removed. Additionally, this case shows significantly reduced particle spread (bottom panel of Fig. 5c) and fewer particles escaping past the filter (top panel of Fig. 5c) than the 5°C temperature difference case (Fig. 5d). The 10°C temperature difference case corresponds to an air temperature of 20°C, which would still allow the musicians to be comfortable while reducing the risk of aerosol accumulation and spread. The filter modelled here has a surface area of 40 × 40 cm^2^, but we expect a filter with a smaller surface area would exhibit a larger change in filtration efficiency when the ambient temperature is changed, as it would provide a smaller target for aerosol particles and would therefore require stronger directional flow to transport them into the filter.

## Conclusions

Through a collaboration with 16 musicians from the Minnesota Orchestra and using multiple measurement techniques and numerical simulations, we conducted a comprehensive assessment of airborne transmission risks during the play of various wind instruments on stage at Orchestra Hall. In general, the flow and aerosol influence zones generated by each instrument are confined to 30 cm from the instrument outlet. The thermal plume generated due to the temperature difference between the human body and the ambient air dominates the flow outside of the influence zone, enhancing mixing and aerosol dispersal. We also found that the outlet flow and aerosol concentration vary with changes in music amplitude, pitch, and note duration, with different relationships observed for each instrument depending on geometry and playing technique. Finally, we tested multiple risk mitigation techniques, including covering the outlets of high-risk instruments (e.g., trumpet) with speaker fabric and placing air filters near the instrument outlets to reduce local aerosol concentration and minimize aerosol spread. Increasing the number of mask layers reduces the aerosol concentration while also deteriorating the sound quality. We show that an air filter placed above the instrument outlet can achieve high efficiency of aerosol reduction even under the lowest flow rate setting (needed to minimize detrimental influence of the filter sound on instrument play) by taking advantage of the upward motion of the human thermal plume that directs particles into the filter. Such filter efficiency can be further increased by raising the temperature difference between the musician body and the ambient air to enhance the effect of thermal plume driving aerosols upward.

Based on our findings, we recommend using masks to cover the bells of the instruments that emit the most aerosols (trumpet, oboe, trombone, bass trombone, and clarinet^15^). The mask should be constructed from a single layer of fabric with low porosity to substantially cut down the aerosol emissions while also minimizing its impact on sound quality. In addition, air filters should be placed above the instrument outlets to minimize aerosol spread and accumulation on stage. To maximize the effectiveness of the filters, we recommend the temperature on stage be reduced to the minimum temperature that is comfortable to the musicians. Furthermore, the recommended 6-foot spacing between musicians should be still be maintained, as they will be breathing and talking when not playing their instruments. Masks should also be worn during breaks in music play.

Note that the findings of the current study are based on experiments conducted at Orchestra Hall in Minneapolis, Minnesota, USA. Variability in ventilation between venues may influence the results, though we expect the thermal body plume flow will still be stronger than ventilation flow in most large concert halls with high ceilings and ventilation inlets located high above or far away from the performance stage. Ten wind instruments were investigated in the current study. Though orchestras may have additional wind instruments, the physical explanations provided here can be used to estimate the influence of any instrument. Finally, we observe significant variabilities between different musicians playing the same instrument, even within the current study. Individual playing styles and instrument setups (e.g., reed stiffness, etc.) may affect some of our results, though we expect they will not influence our main conclusions. In summary, with appropriate precautions taken, our results indicate that musical instrument performance can be undertaken without significant additional risk.

## Methods

### Participants and involved instruments

Sixteen healthy musicians from Minnesota Orchestra aged between 35 and 60 participated in the experiments. Involved musical instruments include trumpet, trombone, bass trombone, French horn, tuba, flute, bassoon, oboe, clarinet, and bass clarinet, which covers nearly all types of wind instruments used in an orchestra. Among them, six of the instruments (trumpet, French horn, flute, bassoon, oboe, and clarinet) are played by two musicians. During the experiment, each musician was asked to play the B-flat major scale at three dynamic levels: *pp* (soft), *mf* (medium), and *ff* (loud). A metronome set to 78 beats per minute was used to maintain consistency, and the musicians were instructed to play two notes per beat (eighth notes). The scale was only performed at the medium dynamic level for the aerosol measurements, as He et al.^15^ (2021) showed that the difference between aerosol emissions at different dynamic levels is relatively small. Additionally, the musicians were asked to prepare two minutes of a musical piece to play (designated as “free play” in the text) to enable correlation between flow speed, aerosol emission, and musical variations. The pieces chosen by the musicians are summarized in Supplementary Table 1.

### Experimental setup

The experiments were conducted on stage at the Minnesota Orchestra Hall. Each experiment lasted 90 minutes, with multiple different measurements conducted including anemometer flow sensing, schlieren flow visualization, and aerosol measurements using aerodynamic particle sizer (APS) and laser sheet and digital inline holography (DIH). Each measurement and data analysis technique is described in more detail below.

### Anemometer flow measurements

A handheld hot-wire anemometer (testo 405i) is used to measure the velocity of the flow coming out from different instruments at different locations. The anemometer is held directly at the instrument outlet, then at 10 cm downstream and 20 cm downstream if flow is still detected. At the outlet, the scales at the three dynamic levels are recorded along with the two minutes of free play. At downstream locations, only the loudest dynamic level scale (strongest flow speed) is measured. After the measurements, the temporal results for the scales are used to calculate the mean velocity and standard deviation under different play conditions and at different locations. The free play time series is used to explore the correlation with the music.

### Schlieren flow measurements

A double pass single mirror schlieren system is used to visualize and quantify the flow fields generated by the musical instruments. A 0.4 m diameter, 2.5 m focal length concave (focusing) mirror is illuminated with a point light source produced by a fiber coupled LED light, placed at a distance of 5 m perpendicular to the mirror plane. The mirror forms an image of the same size at the same distance away from the mirror, but with a lateral displacement proportional to the position of the light. A knife edge is positioned at the focal spot of the mirror to block a portion of the beam and thus enhance the contrast of any light that gets deflected due to variations in index of refraction caused by density/temperature variations within the light path. The light from the schlieren system is captured on a Sony AR 7 II camera equipped with a 200 mm f/2.8 lens imaging at 120 frames per second over 1080 × 720 pixels at a resolution of 0.86 mm/pixel. The musician is seated in front of the mirror with the outlet of the instrument positioned at the edge of the mirror. Background flow is first recorded with the musician seated and holding the instrument in playing position without playing for 60 s. The musician then begins to play and flow from the instrument is captured in the field of view of the camera. Using the recorded intensity patterns, the flow velocity is extracted through correlation-based velocimetry^22,36^. Sample enhanced videos are included in Supplementary Videos 1-5.

### Aerodynamic particle sizer measurements

An aerodynamic particle sizer (APS, TSI model 3321) is used to measure the aerosol concentration emitted by the musicians during instrument play. The APS is capable of capturing particles ranging in size from 0.5 μm to 20 μm. A plastic Y-funnel is used to collect particles, as described in He et al.^15^ (2021). A 50-cm long silicon tube is used to connect the funnel to the APS. The inner diameter of the tube matches the inlet of the APS of 1.2 cm. The funnel is positioned at different locations around the instrument to measure the aerosol dispersion, including directly at the outlet, at different angles relative to the flow, and at different downstream distances.

### Laser sheet aerosol measurements

A 2 W green laser (532 nm wavelength) is projected into a sheet using cylindrical lenses to illuminate aerosol particles in the field of view, where they are captured by a Nikon D610 digital camera with a 50mm Nikkor lens (f/1.2). Videos are captured at 30 frames per second with a shutter speed of 1/30” and ISO 3200. The laser setup as well as the camera are mounted in fixed positions on an L-shaped mounting plate so that the approximately 12 cm × 12 cm field of view is completely covered by the light sheet. The mounting plate itself is fixed to a tripod, allowing adjustments to different angles and heights. A box with a slit placed over the laser and lenses is used to minimize scattered light from reaching the camera. The participant is seated in their natural playing position and the setup is adjusted with the center axis of the examined opening of the instrument facing the camera and perpendicular to the light sheet, as close to the light sheet as possible while minimizing reflections from the instrument (∼3 cm). At the beginning and end of each recording a 10 s segment without any instrument play is recorded; the first 10 s are used to gather information about the background dust level at the orchestra stage while the last 10 s are used to observe aerosols exiting the instrument with a delay. During the recordings, the musicians are seated and instructed to play naturally, while limiting motion of the instrument as much as possible. The videos are analyzed by computing a maximum intensity image for each second (30 frames), then manually counting the particles. This data was only collected for instruments that emit particles at higher concentrations (i.e., trumpet, clarinet). Sample maximum intensity images are shown in Supplementary Fig. 10. These images are used to supplement the APS data and simulations shown in Fig. 2.

### Digital inline holography aerosol measurements

Digital inline holography (DIH), i.e., a high-resolution 3D imaging system, is employed to carry out real time counting and size and shape measurements of the emitted particles with the size ranging from sub-micron to tens of microns. The DIH uses a collimated laser illumination to capture the patterns generated from interference between the light scattered by the particles in the sample volume and un-scattered part of the light, referred to as holograms hereafter^37^. The DIH system uses a Pointgrey grasshopper CCD camera with a sensor size of 3376-pixel x 2704-pixel and a Thorlabs 532 nm continuous laser. With a 4X magnification imaging lens, the DIH system has a pixel resolution of 0.7 µm and the sample volume is 2.3 mm × 1.8 mm × 10 mm. An in-house developed hologram processing software which combines hologram acquisition, enhancement, and automatic selection of frames with emitted particles is used to collect hologram sequence. The software is running on a Tensorbook laptop connected with the camera through USB 3.0 interface. The laptop automatically collects the hologram data and generates the numbered list of holograms with particles. After data collection, the holograms with particles are numerically reconstructed and thresholded for measuring particle concentration, and size and shape distribution. During the experiments, the participants are instructed to directly point the outlet of the instruments to a funnel connected to the measurement volume of the DIH system through a 50-cm long silicon tube. The time of measurement is two minutes for each musician. The resulting size and shape histograms of the captured aerosols are presented in Supplementary Fig. 11. This data is used to supplement the particle size measurements from the APS.

### Thermal imaging based human model

In order to accurately capture the impact of realistic orchestra playing environments on aerosol emission and transmission, a multiscale flow domain including a virtual musician and instrument is used in the simulations. Thermal images of a healthy subject were recorded during playing the trumpet. The acquired images were used to reconstruct the 3D virtual manikin and instrument. In this model, the body is composed of cube segments with dimensions based on trumpet player 1 (see Supplementary Fig. 8). The response of human temperature to environment and water vapor flux through the skin surface are ignored during the simulation time, and direct heat conduction in adjacent segments is neglected. Fixed temperature distribution boundary conditions on the human body are used. The average body temperature from the thermal images was use for the boundary condition due to the relatively small differences in skin temperature across different parts of the body^38^. Long wave radiative heat transfer is also coupled with a convective heat transfer analysis with CFD. The bells of the instruments played by each musician are modelled and positioned in front of the virtual manikin to realistically model the outlet air jet conditions during instrument play (see Supplementary Fig. 8). A particle emission source is created at the instrument outlet, the rate of which is obtained through measurements obtained using the aerodynamic particle sizer.

### CFD set-up

The 3D model was imported into OpenFOAM^®^ software for CFD analysis. The computational domain for the following simulation is a rectangular chamber of 8 × 8 × 10 m^3^. The musician sits on the bottom x-z surface (see Supplementary Fig. 8). A no-slip wall boundary condition is applied at the bottom floor, human body and instrument surface, and an outlet pressure boundary condition at the top surface (y direction). The remaining boundaries are treated as symmetry plane boundary conditions in the two horizontal directions (x and z direction). Temperature is specified at the bottom floor and human body (head, trunk, arms, hands, legs, and feet) and zero gradient boundary conditions at the top surface. A mass flow rate boundary condition is assigned at the instrument bell to simulate the effect of the instrument outlet air jet on the local flow field and the aerosol dispersion over the entire sample volume. The musician is oriented in the *x* direction. The implicit unsteady shear stress transport k-ω turbulence model is used with low Reynolds number modification to model the flow turbulence, which has been previously used in respiration studies^39,40^.

Simulations are conducted based on the OpenFOAM-6 platform, with the Eulerian-Lagrangian framework for the gas-liquid phase simulation. Air flow is calculated using a compressible solver to model the buoyant forces; particle movement is calculated by solving Newton’s 2^nd^ Law.

The carrier phase (air flow) is modelled using the following equations:

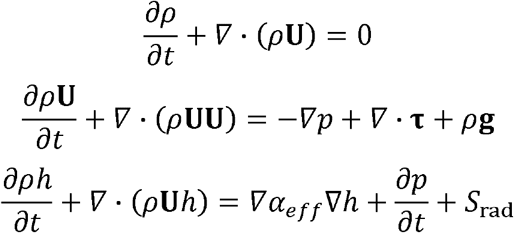

where ρ is fluid density, *U* represents the velocity, *g*=9.81 m/s^2^ is the acceleration due to gravity, *p* is the pressure, *h* is the enthalpy, τ is the stress tensor, α_eff_ is the effective thermal diffusivity, and *S*_rad_ is a source term for the radiative heat transfer. Considering the dilute particle concentration in the present study, the droplet induced mass, momentum, and energy transfer are ignored.

The one-way coupled Euler-Lagrange approach is applied to predict the dispersion and deposition of each aerosol generated during instrument play. Particles are assumed to be spherical and liquid and particle-particle interactions are ignored. The translational motion of each particle is governed by the force balance equation, i.e., the Maxey-Riley equation. To determine the particle velocity *u*_iP_, and position *x*_iP_, the force balance equation is solved for each particle, which is given by:

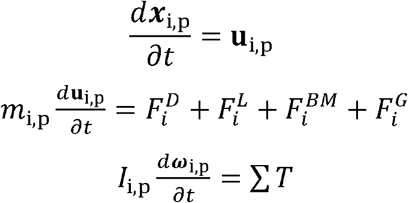

where *i* is the particle ID, *u*_p_ is the particle velocity, *m*_p_ is the particle mass, *F*^D^ represents the drag force^41^, *F*^L^ is the Saffman lift force^42^, *F*^G^ is the gravitational force, and *F*^BM^ is the Brownian motion induced force^43^. The Kelvin effect is not considered, because our experimental data indicates that the particle number density is too low for collision and particle sizes are too low for break up.

### Grid generation

ICEM 18.0 is used to generate the hex-core meshes. Ten layers of mesh is applied over the human body model to resolve the boundary layer. The first layer distance is set as 0.005 m so that the y^+^ around human body is in the range of the wall function usage requirement^44^. The cell size of the mesh for different cases ranges from 7 million 10 million.

### Numerical validation

The ability of the current numerical methodology to predict the human thermal environment has been validated against experimental data. The velocity induced by the human thermal plume was measured at locations 5 cm, 10 cm, 20 cm, 40 cm, 80 cm above the head. The simulation result probing at same location agrees well with the experiment data, as shown in Supplementary Fig. 9. The maximum velocity above the human head is 0.22 m/s at 0.5 m.

## Data Availability

All data is available in the main text or the supplementary materials. All data, code, and materials are hosted at the Data Repository for the University of Minnesota.

## Acknowledgments

This work was supported by the School of Medicine of the University of Minnesota. The authors would like to thank Dr. Kevin Mallery for his assistance with experiment preparation. We would also like to thank Prof. David Y.H. Pui for equipment support, his postdocs Dr. Qisheng Ou and Dr. Seong Chan Kim and graduate student Dongbin Kwak for their assistance in the equipment calibration. In addition, we thank Joel Mooney and the other Minnesota Orchestra staff members who assisted with the experiments, as well as the 16 musicians from the Minnesota Orchestra for their participation in the experiments. Finally, we thank Prof. Jon Hallberg for connecting us with the Minnesota Orchestra.

## Author Contributions

A.A. and R.H. contributed equally. A.A., R.H., B.G., S.S., S.S.K., M.T., R.G.P., and J.H. conducted experiments; R.H. and C.W. conducted numerical simulations; A.A., R.H., and S.S. performed data analysis; A.A. wrote the manuscript; J.H. and S.S.K. revised the manuscript; M.W. provided professional interpretation and organized experiments; J.H. supervised the project.

## Declaration of competing interests

MW is employed by the Minnesota Orchestra. No other relationships or activities that could appear to have influenced the submitted work.

## Notes

### Author Declarations

The University of Minnesota provided IRB approval for this study (HSIRB STUDY00009795)

