## Supplementary Information for "Risk Assessment and Mitigation of Airborne Disease Transmission in Orchestral Wind Instrument Performance"

**Affiliations:**

### Supplementary Figures

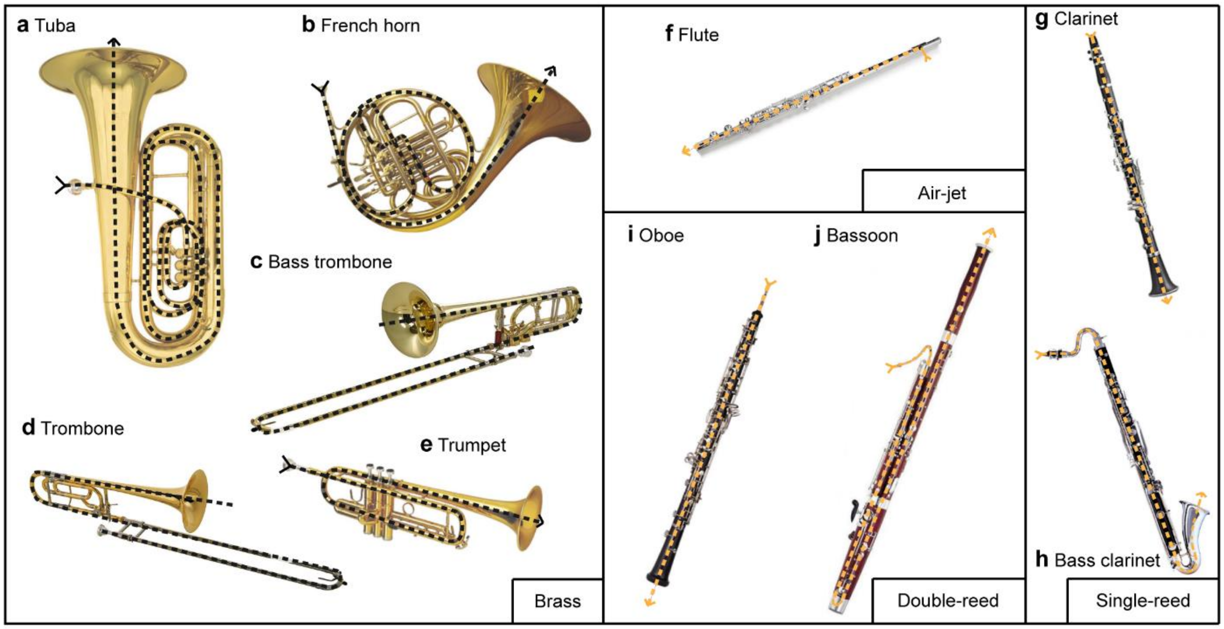


**Supplementary Figure 1 | Images of the ten instruments investigated.** Brass instruments include (**a**) tuba, (**b**) French horn, (**c**) bass trombone, (**d**) trombone, and (**e**) trumpet. The air-jet woodwind is (**f**) flute. Single-reed woodwinds include (**g**) clarinet and (**h**) bass clarinet. Double-reed woodwinds include (**i**) oboe and (**j**) bassoon. The dashed line marks the main flow path in each instrument.

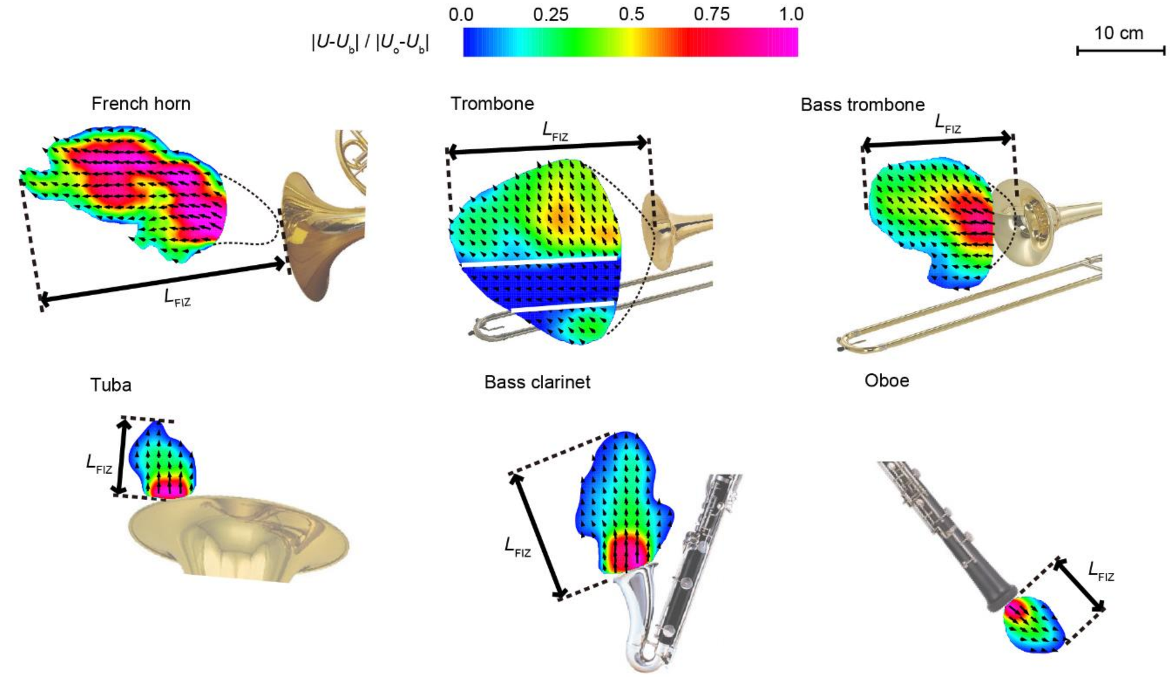


**Supplementary Figure 2 | Flow influence zones of all instruments.** Mean velocity fields showing the flow influence zones of all instruments not included in the main text. The near-field velocity fields for the French horn, trombone, and bass trombone are not available due to the presence of the musician's hand in or near the bell (these regions have been removed in the figures). The region of the trombone velocity field demarcated by white lines indicates the region blocked by the slide.

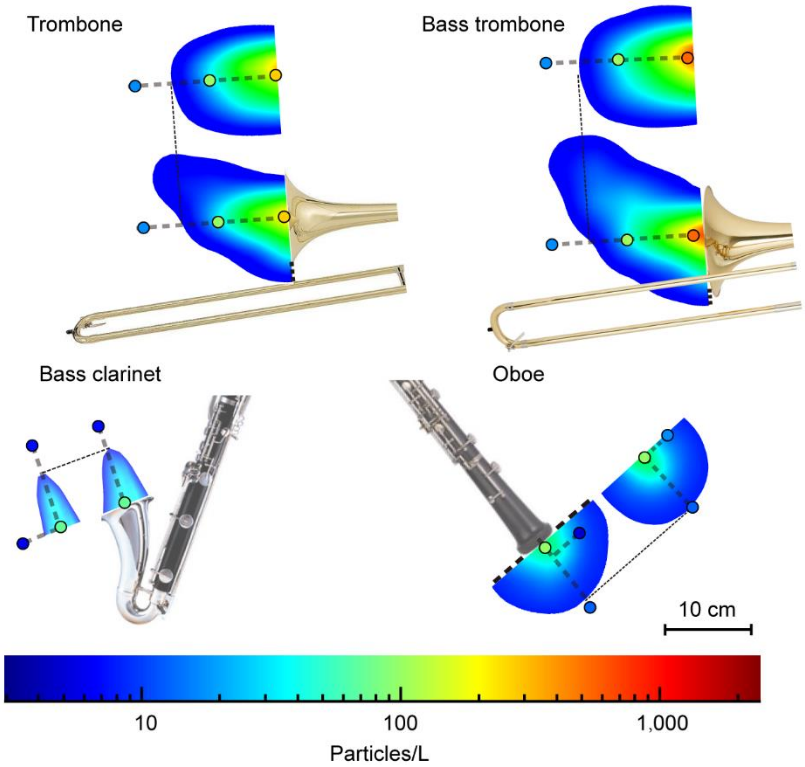


**Supplementary Figure 3 | Aerosol influence zones of all instruments.** The aerosol concentration throughout cross-sections of the instrument aerosol influence zone for all instruments not shown in the main text. The dots indicate the locations and values of experimental APS measurements. Note that French horn (played with the musician's hand in the bell) and tuba are not included because they both emit aerosol concentrations at the background level based on APS measurements.

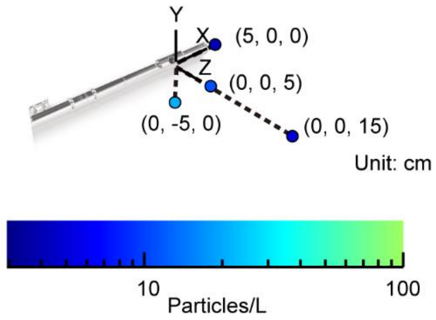


**Supplementary Figure 4 | Aerosol concentration from flute embouchure.** The aerosol concentration emitted from the flute embouchure measured using the APS. This region was not simulated due to the complexity of the flow from the mouth over the mouthpiece. Previous studies (Becher et al. 2020 and Kähler & Hain 2020) have suggested that the flute is one of the riskiest instruments due to the large region influenced by the flow from the embouchure, but this data shows that the aerosol concentration in this region is extremely low. The background particle concentration in Orchestra Hall is 14.4 particles/L.

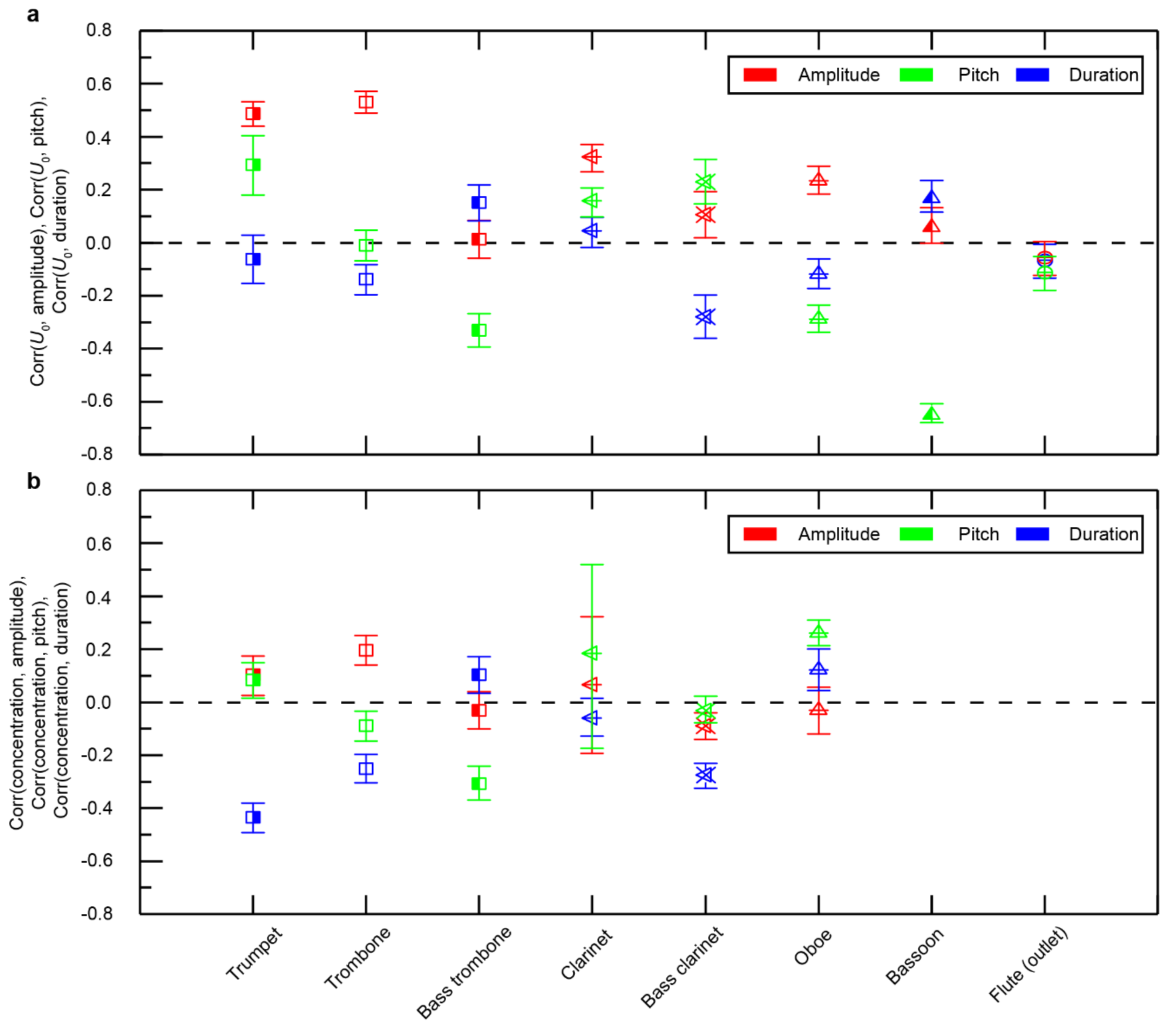


**Supplementary Figure 5 | Correlations between music, flow, and particle concentration.** Correlation coefficients for all instruments between amplitude, pitch, duration, and (**a**) flow speed, or (**b**) particle concentration. The error bars represent the probable error of each correlation coefficient. Note that correlations with aerosol concentration are not provided for flute and bassoon due to the very low emission level of these instruments (see Fig. 2 and He et al.[15]).

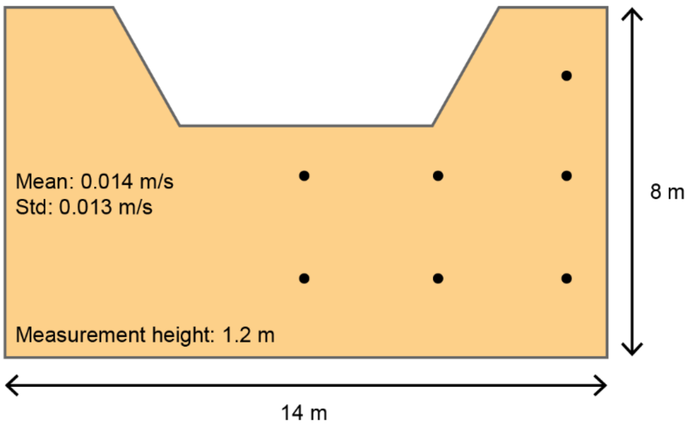


**Supplementary Figure 6 | Ventilation on the Orchestra Hall stage.** Map of the measurement locations of ventilation flow speed, $U_{\mathrm{vent}}$, on the upper deck of the stage in Orchestra Hall. Only the upper deck is included, as that is where the wind instruments are seated during performances.

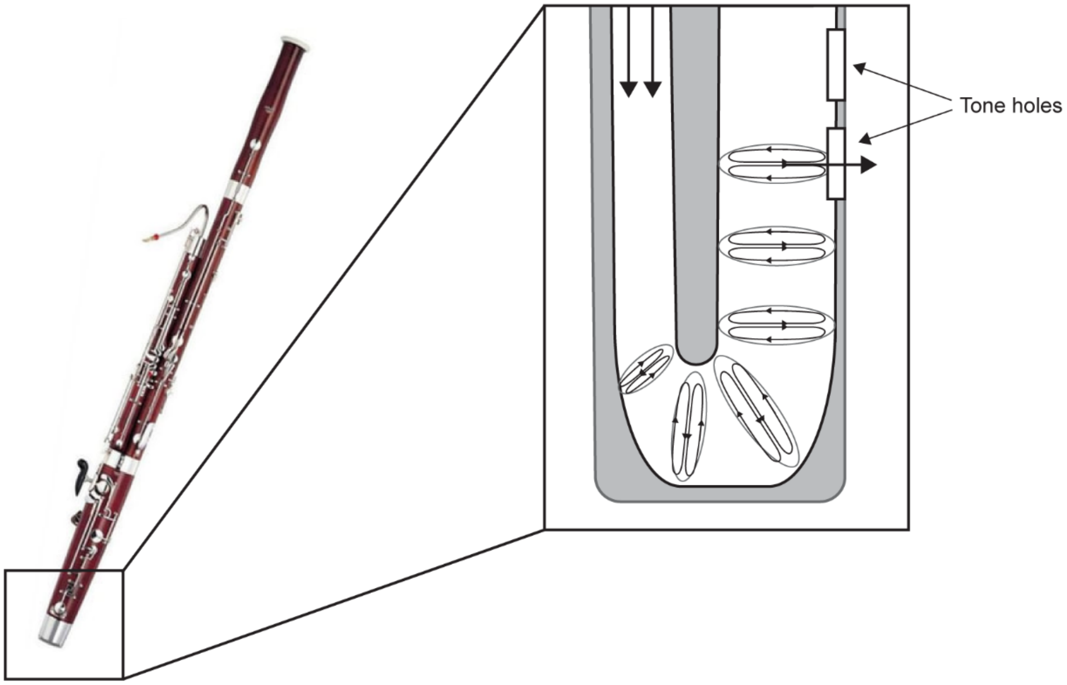


**Supplementary Figure 7 | Dean flow inside the bassoon bore.** Schematic showing the development of a secondary flow (Dean flow) that develops within the 180° bend at the bottom of the bassoon. Dean flow persists downstream of the bend, pushing air out of the tone holes.

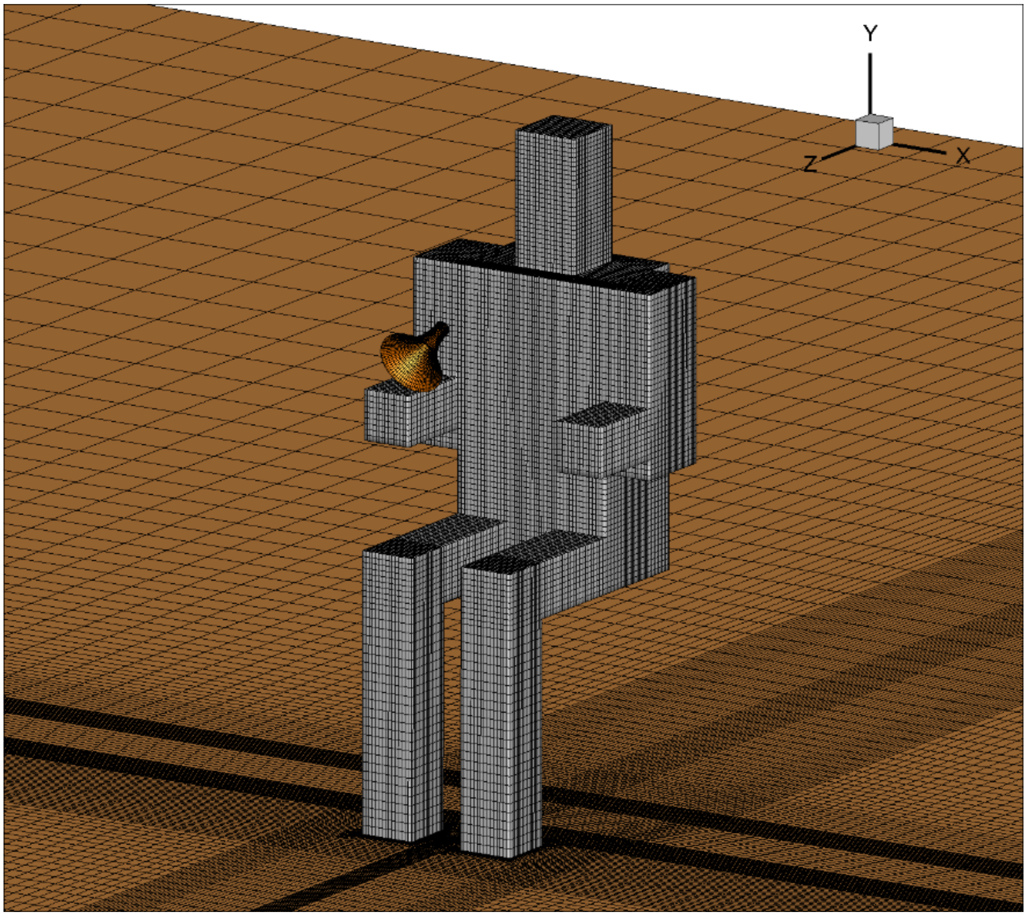


**Supplementary Figure 8 | 3D virtual manikin.** Cube-based human body model used for all CFD simulations. The dimensions are based on the dimensions of trumpet player 1.

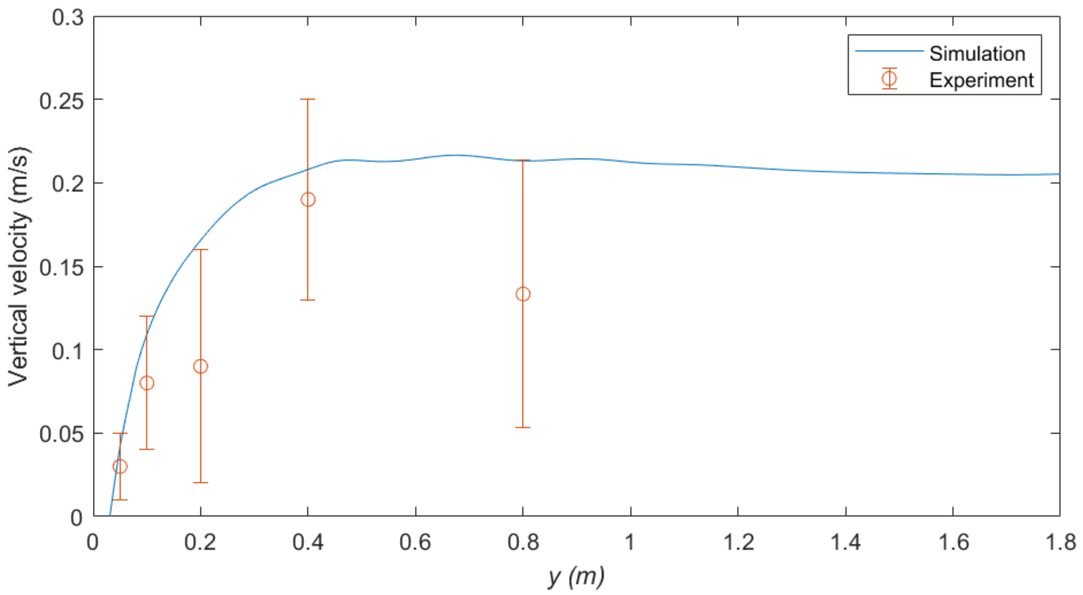


**Supplementary Figure 9 | Numerical validation.** Results of numerical validation test of the velocity above the human head induced by the thermal plume.

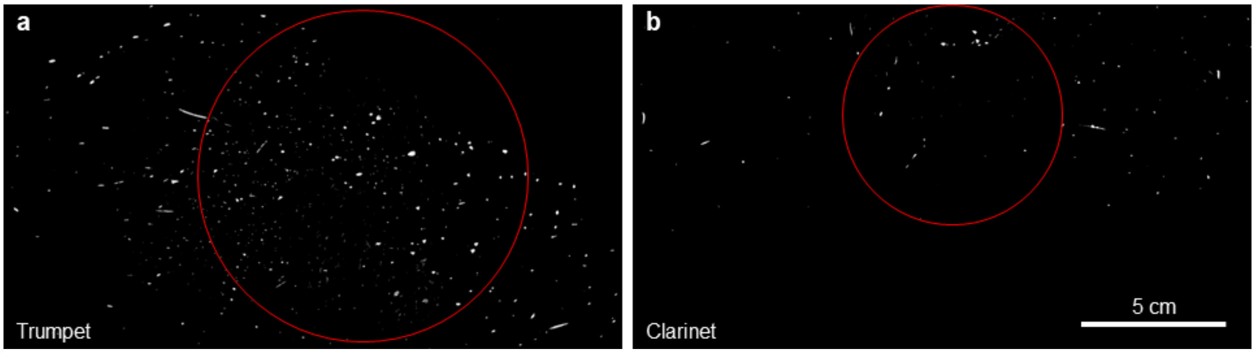


**Supplementary Figure 10 | Laser sheet images.** Maximum intensity projection of the images of aerosols emitted during one minute of scale play from (**a**) a trumpet and (**b**) a clarinet as they pass through the laser sheet. The aerosols can be seen as small dots. The red circles indicate the locations of the instrument bells.

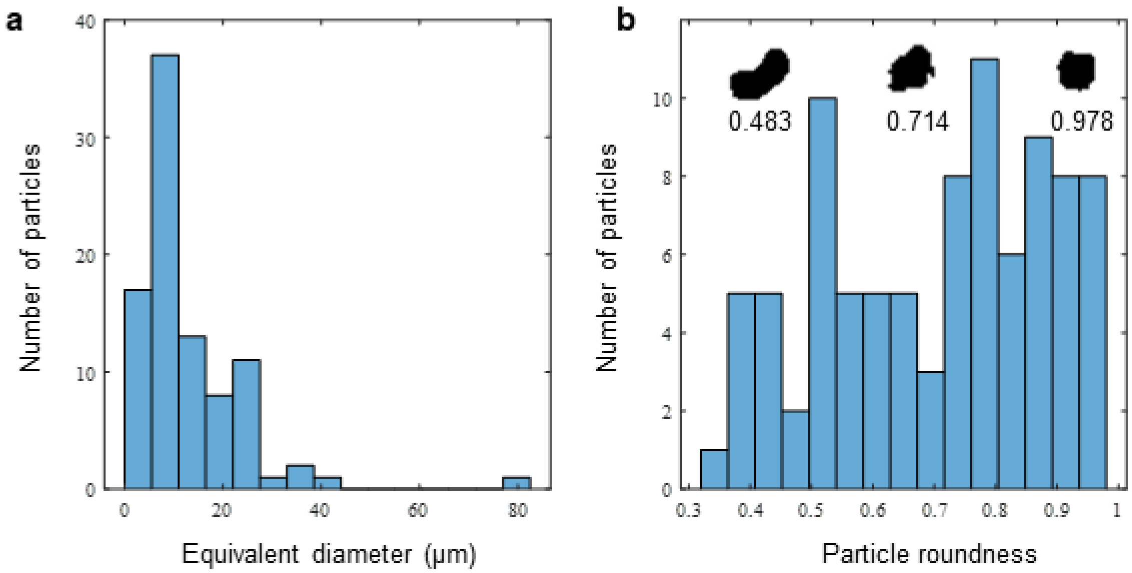


**Supplementary Figure 11 | Aerosol properties.** Distribution of (**a**) equivalent diameter and (**b**) roundness of aerosols emitted from all musicians, measured using digital inline holography (DIH).

**Supplementary Tables**

**Supplementary Table 1 | Music piece performed by each musician.**

| Instrument | Piece | Composer |
| --- | --- | --- |
| Tuba | The Ride of the Valkyries | Wagner |
| | Die Meistersinger | Wagner |
| French horn 1 | Horn Concerto #2 in E flat | Mozart |
| French horn 2 | Horn Concerto #1 | Mozart |
| Bass trombone | Sarabande | Bach |
| | The Ride of the Valkyries | Wagner |
| | The Creation | Haydn |
| | Symphony no. 9 | Beethoven |
| Trombone | Requiem Tuba Mirum | Mozart |
| | The Ride of the Valkyries | Wagner |
| Trumpet 1 | Légende | Enescu |
| | Indivisible | Heitzeg |
| Trumpet 2 | Symphony no. 8 | Mahler |
| | Pictures at an Exhibition: Promenade | Mussorgsky/Ravel |
| Flute 1 | Daphnis et Chloe | Ravel |
| Flute 2 | Syrinx | Debussy |
| Clarinet 1 | Time Study no. 1 | Stulz |
| Clarinet 2 | Octet in F major | Schubert |
| Bass clarinet | Etude no. 1 of 32 | Rose |
| Oboe 1 | Gabriel’s Oboe | Morricone |
| Oboe 2 | Fantasia no. 4 in B-flat major | Telemann |
| Bassoon 1 | Bassoon Concerto | Mozart |
| Bassoon 2 | Bassoon Concerto | Mozart |

**Supplementary Table 2 | Geometry of the 10 instruments tested in the study.**

| Instrument | Tube length (m) | Bore diameter (mm) | Bell diameter (mm) |
|---|---|---|---|
| Tuba | 4.9 | 11 | 480 |
| French horn | 3.7 | 11 | 305 |
| Bass trombone | 2.75 | 14.3 | 247 |
| Trombone | 2.7 | 13.6 | 212 |
| Trumpet | 1.4 | 10.7 | 114 |
| Flute | 0.66 | 19 | -- |
| Clarinet | 0.66 | 12.7 | 76 |
| Bass clarinet | 0.94 | 25.4 | 130 |
| Oboe | 0.64 | 4.5-43 (cone) | 43 |
| Bassoon | 2.56 | 4-40 (cone) | 40 |

Instrument geometries from the following references: Berkopec, 2013; Bucur, 2019; Campbell, Greated, & Myers, 2004; Thompson, 2010.

**Supplementary Videos**

**Supplementary Video 1 | Trumpet air flow.** Sample section of enhanced schlieren video showing air flow from the trumpet.

**Supplementary Video 2 | Clarinet air flow.** Sample section of enhanced schlieren video showing air flow from the clarinet.

**Supplementary Video 3 | Flute air flow.** Sample section of enhanced schlieren video showing air flow from the flute.

**Supplementary Video 4 | Bassoon air flow.** Sample section of enhanced schlieren video showing air flow from the bassoon.

**Supplementary Video 5 | Bass clarinet air flow.** Sample section of enhanced schlieren video showing air flow from the bass clarinet.

**Supplementary Video 6 | Trumpet aerosol transport.** Sample video showing the simulated aerosol particles emitted from the trumpet.

**Supplementary Video 7 | Trumpet combined signals.** Video with trumpet music audio, amplitude, pitch, schlieren video, velocity from anemometer, and aerosol concentration from APS. All signals have been synchronized based on the music audio.

**Supplementary Video 8 | Clarinet combined signals.** Video with clarinet music audio, amplitude, pitch, schlieren video, velocity from anemometer, and aerosol concentration from APS. All signals have been synchronized based on the music audio.

**Supplementary Video 9 | Oboe combined signals.** Video with oboe music audio, amplitude, pitch, schlieren video, velocity from anemometer, and aerosol concentration from APS. All signals have been synchronized based on the music audio.

**Supplementary Video 10 | Bassoon combined signals.** Video with bassoon music audio, amplitude, pitch, schlieren video, velocity from anemometer, and aerosol concentration from APS. All signals have been synchronized based on the music audio.

**Supplementary Video 11 | Trumpet music play without filter.** Video showing simulated aerosol particles and corresponding $\bar{I}_{\text{risk}}$ for a sequence of trumpet music play without a filter. The difference between the ambient and human body temperature is 10°C.

**Supplementary Video 12 | Trumpet play with filter in front.** Video showing simulated aerosol particles and corresponding $\bar{I}_{\text{risk}}$ for a sequence of trumpet music play with a filter placed on the floor in front of the musician. The difference between the ambient and human body temperature is 10°C.

**Supplementary Video 13 | Trumpet play with filter above.** Video showing simulated aerosol particles and corresponding $\bar{I}_{\text{risk}}$ for a sequence of trumpet music play with a filter placed above the instrument outlet. The difference between the ambient and human body temperature is 10°C.

**Supplementary Video 14 | Trumpet play with filter above, 5°C.** Video showing simulated aerosol particles and corresponding $\bar{I}_{\text{risk}}$ for a sequence of trumpet music play with a filter placed above the instrument outlet. The difference between the ambient and human body temperature is 5°C.
